# Impact of *BRCA* mutation status on tumor infiltrating lymphocytes (TILs), response to treatment, and prognosis in breast cancer patients treated with neoadjuvant chemotherapy

**DOI:** 10.1101/2020.09.27.20202515

**Authors:** Beatriz Grandal, Clémence Evrevin, Enora Laas, Isabelle Jardin, Sonia Rozette, Lucie Laot, Elise Dumas, Florence Coussy, Jean-Yves Pierga, Etienne Brain, Claire Saule, Dominique Stoppa-Lyonnet, Sophie Frank, Claire Sénéchal, Marick Lae, Diane De Croze, Guillaume Bataillon, Julien Guerin, Fabien Reyal, Anne-Sophie Hamy

**Affiliations:** Department of Surgery, Institut Curie, 26 rue d’Ulm, 75005 Paris, France; Department of Oncology, Institut Curie, 26 rue d’Ulm, 75005 Paris, France; Department of Oncology, Centre René Huguenin – Institut Curie, 35 rue Dailly, 92210 St Cloud, France; Department of Genetics, Institut Curie, 26 rue d’Ulm, 75005 Paris, France INSERM U830, Institut Curie Paris, Paris, France; Department of Genetics, Institut Bergonié, 229 Cours de l’Argonne, 33000 Bordeaux, France; Department of Pathology, Centre René Huguenin - Institut Curie, 35 rue Dailly, 92210 St Cloud, France; Department of Pathology, Centre Henri Becquerel, INSERM U1245, uniRouen, University of Normandie, Rouen, France; Department of Pathology, Institut Curie, 26 rue d’Ulm, 75005 Paris, France; Data Office, Institut Curie, 25 rue d’Ulm, 75005 Paris, France; Residual Tumor & Response to Treatment Laboratory, RT2Lab, translational Research Department, INSERM, U932 Immunity and Cancer, Institut Curie, 26 rue d’Ulm, Paris, France

**Keywords:** BRCA, TILs, pCR, NAC, immunotherapy

## Abstract

**Introduction:** Five to 10% of breast cancers (BCs) occur in a genetic predisposition context (mainly BRCA pathogenic variant). Nevertheless, little is known about immune tumor infiltration, response to neoadjuvant chemotherapy (NAC), pathologic complete response (pCR) and adverse events according to *BRCA* status.

**Material and methods:** Out of 1199 invasive BC patients treated with NAC between 2002 and 2012, we identified 267 patients tested for a germline *BRCA* pathogenic variant. We evaluated pre-NAC and post-NAC immune infiltration (TILs). Response to chemotherapy was assessed by pCR rates. Association of clinical and pathological factors with TILs, pCR and survival was assessed by univariate and multivariate analyses.

**Results:** Among 1199 BC patients: 46 were *BRCA*-deficient and 221 *BRCA-*proficient or wild type (WT). At NAC completion, pCR was observed in 84/266 (31%) patients and pCR rates were significantly higher in *BRCA-*deficient BC (*p=* 0.001), and this association remained statistically significant only in the luminal BC subtype (*p=* 0.006). The interaction test between BC subtype and *BRCA* status was nearly significant (*P*_*interaction*_=0.056). Pre and post-NAC TILs were not significantly different between *BRCA-*deficient and *BRCA-*proficient carriers; however, in the luminal BC group, post-NAC TILs were significantly higher in *BRCA-*deficient BC. Survival analysis were not different between *BRCA-*carriers and non-carriers.

**Conclusion:** **BRCA** mutation status is associated with higher pCR rates and post-NAC TILs in patients with luminal BC. *BRCA*-carriers with luminal BCs may represent a subset of patients deriving higher benefit from NAC. Second line therapies, including immunotherapy after NAC, could be of interest in non-responders to NAC.

**Translational relevance:** High lymphocytic infiltration (TILs) seem to reflect favorable host antitumor immune responses. In breast cancer, the variation of TILs before and after neoadjuvant chemotherapy (NAC) according to *BRCA* status has been poorly described. Little data is available on their value after treatment. We investigated TIL levels before and after NAC and response to treatment in 267 paired biopsy and surgical specimens.

In our study, luminal BCs were associated with pathologic complete response (pCR) and higher TIL levels after chemotherapy completion in patients with *BRCA* pathogenic mutations. Our data supports that (i) NAC should be reconsidered in luminal BCs with *BRCA* pathogenic mutation, (ii) TILs could be a biomarker for response to immune checkpoint blockade in luminal BCs with *BRCA* pathogenic variant who did not achieve a pCR and (iii) exploiting the antitumor immune response in luminal BCs could be an area of active research.

## Introduction

**Neoadjuvant or pre-operative chemotherapy (NAC)** is classically administered to patients with inflammatory or locally advanced breast cancer (BC). Beyond increasing breast-conserving surgery rates (1), it also serves as an *in vivo* chemosensitivity test and the analysis of residual tumor burden may help understanding treatment resistance mechanisms (2). In addition, it helps refining the prognosis of patients after NAC, as pathological complete response (pCR) after NAC is associated with a better long term survival (1,3).

Nearly 5% of breast cancers occur in a context of **genetic predisposition**, mostly represented by monoallelic pathogenic variants of *BRCA1, BRCA2* or *PALB2* genes (4). Patients with loss-of-function of the BRCA1 or 2 proteins have a higher cumulated breast cancer risk, with a cumulated life time risk at eighty years old of 72% (*BRCA1*) and 69% (*BRCA2*)(5). The peak incidence for *BRCA1* mutation carriers occurs between 41 and 50 years old (28.3 per 1000 person-years), whereas it occurs ten years later for *BRCA2* mutation carriers (30.6 per 1000 person-years between 51 and 60) (5). *BRCA1* and *BRCA2* are tumor-suppressor genes that code for proteins involved in homologous recombination (HR) repair. HR deficiency (HRD) occurs when the second allele is inactivated by allelic deletion (often detected by LOH), genic alteration or promoter methylation (for *BRCA1* only). Biallelic *BRCA1/2* inactivation results in genomic instability and theoretically increases the somatic mutational load (6).

Tumors associated with germline or somatic *BRCA1/2* pathogenic mutations **display different patterns when compared with sporadic BCs**. Cancers occurring among *BRCA1* carriers are more frequently classified as medullary (7), whereas histological subtypes among *BRCA2* carriers tend to be more heterogeneous (8). In addition, *BRCA1* carriers are more frequently ER-negative, PR-negative and lack *HER2* amplification (*i*.*e*. display a triple negative (TNBCs) phenotype (9))- whereas in *BRCA2* carriers, a similar prevalence of ER-positive tumors has been described when compared with sporadic controls (10–13).

Most of patients with TNBCs receive **chemotherapy** (14,15). Due to the alteration of BRCA1 and BRCA2 proteins in tumor cells, *BRCA*-mutated cells are unable to properly repair double-strand breaks, classically induced by DNA-alkylating agents (16). Hence, *BRCA* deficiency has sometimes been associated with a higher sensitivity to platinum agents when compared to other types of neoadjuvant chemotherapy regimens (17–19). However, the effectiveness of standard NAC in all BC subtypes associated with *BRCA* pathogenic variants compared to controls has been poorly explored so far.

The role of **tumor infiltrating lymphocytes (TILs)** in BC has been extensively studied over the last decade. High levels of TILs before NAC are associated with higher pCR rates and better survival, especially for TNBC and *HER2*-positive BCs (20,21). However, despite a growing interest in the field of immunity and oncology, characterization and quantification of TILs across all BC subtypes according to *BRCA* status has not been extensively described. Similarly, no study has evaluated so far, the evolution of immune infiltration after NAC according to *BRCA* status.

The objective of the current study is to determine if pre and post-NAC TILs, chemosensitivity and prognosis differ according to *BRCA* status in a cohort of BC patients treated with NAC.

## Material and methods

### Patients and Tumors

The study was performed on a retrospective institutional cohort of 1199 female patients with T1-T3NxM0 invasive BC (NEOREP Cohort, CNIL declaration number 1547270) treated with NAC at Institute Curie (Paris and Saint-Cloud) between 2002 and 2012. The cohort included unifocal, unilateral, non-recurrent, non-metastatic tumors, excluding T4 tumors (inflammatory, chest wall or skin invasion). Approved by the Breast Cancer Study Group of Institute Curie, the study was conducted according to institutional and ethical rules concerning research on tissue specimens and patients. Informed consent from patients was not required.

Information on family history, clinical characteristics (age; menopausal status; body mass index) and tumor characteristics (clinical tumor stage and grade; histology; clinical nodal status; ER, PR and *HER2* status; BC subtype; mitotic index; Ki67) were retrieved from electronic medical records. All the patients received NAC, and additional treatments were decided according to national guidelines (see **Supplementary material**).

### Tumors samples

In accordance with French national guidelines (22), cases were considered estrogen receptor (ER)-positive or progesterone receptor (PR)-positive if at least 10% of tumor cells expressed estrogen and/or progesterone receptors (ER/PR), and endocrine therapy was prescribed when this threshold was exceeded. *HER2* negative status was defined as 0 or 1 + on immunohistochemistry (IHC) stained tissue section. IHC 2 + scores were subsequently analyzed by fluorescence *in situ* hybridization (FISH) to confirm *HER2* positivity. Pathological BC were classified into subtypes (TNBC, *HER2*-positive, and luminal *HER2*-negative [referred to hereafter as “luminal”]) (see **Supplementary material**).

### TIL levels, pathological complete response and pathological review

TIL levels were evaluated retrospectively for research purposes, by two pathologists (ML and DdC) specialized in breast cancer. TIL levels were assessed on formalin-fixed paraffin-embedded (FFPE) tumor tissue samples from pretreatment core needle biopsies and the corresponding post-NAC surgical specimens, according to the recommendations of the international TILs Working Group before (23) and after NAC (24). TILs were defined as the presence of a mononuclear cell infiltrate (including lymphocytes and plasma cells, excluding polymorphonuclear leukocytes). TILs in direct contact with tumor cells were counted as intra-tumoral TILs (IT TILs) and those in the peri-tumoral areas as stromal TILs (str TILs). They were evaluated both in the stroma and within tumor scar border, after excluding areas around ductal carcinoma *in situ*, tumor zones with necrosis and artifacts, and were scored continuously as the average percentage of stroma area occupied by mononuclear cells. We defined pathological complete response (pCR) as the absence of invasive residual tumor from both the breast and axillary nodes (ypT0/is N0).

### BRCA status

Genetic counseling was offered based on individual or family criteria (see Supplemental material). When constitutional genetic analysis of *BRCA1* and *BRCA2* genes were required, Denaturing High Performance Liquid Chromatography (DHPLC) and Sanger sequencing were performed to search for point alterations, and Quantitative Multiplex Polymerase Chain Reaction of Short Fluorescent (QMPSF) to research large gene rearrangements between 2002 and 2012. In case of previously known pathogenic familial variants, targeted tests were performed.

### Survival endpoints

Relapse-free survival (RFS) was defined as the time from surgery to death, loco-regional recurrence or distant recurrence, whichever occurred first. Overall survival (OS) was defined as the time from surgery to death. For patients for whom none of these events were recorded, data was censored at the time of last known contact. Survival cutoff date analysis was February 1^st^, 2019.

### Statistical analysis

Pre- and post-NAC TIL levels were analyzed as continuous variables. All analyses were performed on the whole population and after stratification by BC subtype. To compare continuous variables among different groups, Wilcoxon-Mann-Whitney test was used for groups including less than 30 patients and for variables displaying multimodal distributions; otherwise, student t-test was used. Association between categorical variables was assessed with chi-square test, or with the Fisher’s exact test if at least one category included less than three patients. In boxplots, lower and upper bars represented the first and third quartile respectively, the medium bar was the median, and whiskers extended to 1.5 times the inter-quartile range. Factors predictive of pCR were introduced in a univariate logistic regression model. Covariates selected for multivariate analysis were those with a *p*-value no greater than 0.1 after univariate analysis. Survival probabilities were estimated by Kaplan-Meyer method, and survival curves were compared with log-rank tests. Hazard ratios (HR) and their 95% confidence intervals (CI) were calculated with the Cox proportional hazard model. Analyses were performed with R software version 3.1.2. Significance threshold was of 5%.

## Results

### Study population and tumors characteristics

The total number of patients included in the neoadjuvant cohort was 1199. Among the whole population, germline *BRCA* pathogenic variant status was available for 267 patients (22.3%), and was not obtained for 932 patients (77.73%, **Supplementary Figs. S1)**. Median age of cohort’s population was 48 years old (range 24-80) and most patients (n=747, 62%) were premenopausal. Median BMI index was 24.74, and 25.8% had direct family history of breast cancer. Patients repartition by subtype was as follows: luminal (n=518, 44%), TNBC (n= 376, 31%), *HER2*-positive (n= 295, 25%).

Patients with available *BRCA* status were significantly different from patients with *BRCA* status unknown. They were younger, had lower body mass index, were more likely to be diagnosed with grade III, TNBC of no specific type (NST), and to receive standard anthracyclines-taxanes containing regimens than patients not screened (p< 0.001) (**Table1, Supplementary Figs. S2)**.

**Table1.** Patients’characteristics among the whole population

| Characteristics | Class | All | BRCA mutation | BRCA wild-type | Not screened | p |
| --- | --- | --- | --- | --- | --- | --- |
| n= |  | 1199(100%) | 46(3.8%) | 221(18.4%) | 932(77.7%) |  |
| Age (mean) |  | 48.6 | 39.5 | 41.7 | 50.6 | <0.01 |
| Menopausal status | pre | 747 (62.8) | 41 (89.1%) | 187 (85.0%) | 519 (56.2%) | <0.01 |
|  | post | 442 (37.2) | 5 (10.9%) | 33 (15.0%) | 404 (43.8%) |  |
| BMI (mean) |  | 24.7 | 22.8 | 23.6 | 25.1 | <0.01 |
| BMI class | [15,19] | 72 (6.0) | 6 (13.3) | 17 (7.7) | 49 (5.3) | <0.01 |
|  | (19,25] | 664 (55.7) | 31 (68.9) | 147 (66.5) | 486 (52.4) |  |
|  | (25,30] | 299 (25.1) | 4 (8.9) | 43 (19.5) | 252 (27.2) |  |
|  | (30,50] | 158 (13.2) | 4 (8.9) | 14 (6.3) | 140 (15.1) |  |
| Family history of BC | no | 887 (74.2) | 12 (26.1%) | 104 (47.7%) | 771 (82.7%) | <0.01 |
|  | yes | 309 (25.8) | 34 (73.9%) | 114 (52.3%) | 161 (17.3%) |  |
| Clinical tumor size | T1 | 70 (5.8%) | 5 (10.9%) | 22 (10.0%) | 43 (4.6%) | <0.01 |
|  | T2 | 798 (66.6%) | 28 (60.9%) | 153 (69.2%) | 617 (66.3%) |  |
|  | T3 | 330 (27.5%) | 13 (28.3%) | 46 (20.8%) | 271 (29.1%) |  |
| Clinical nodal status | N0 | 525 (43.8%) | 17 (37.0%) | 93 (42.1%) | 415 (44.6%) | 0.51 |
|  | N1-N2-N3 | 673 (56.2%) | 29 (63.1%) | 128 (57.9%) | 516 (55.4%) |  |
| Histology | NST | 1062 (90%) | 43 (93.5%) | 213 (96.4%) | 806 (88.3%) | 0.03 |
|  | others | 118 (10%) | 3 (6.5%) | 8 (3.6%) | 108 (11.6%) |  |
| Grade | I-II | 479 (41.4%) | 10 (23.3%) | 76 (34.7%) | 393 (43.9%) | 0.01 |
|  | III | 678 (58.6%) | 33 (76.7%) | 143 (65.3%) | 502 (56.1%) |  |
| Mitotic Index (mean) |  | 25.1 | 30.8 | 25.6 | 24.6 | 0.25 |
| Subtype | luminal | 528 (44.0%) | 15 (32.6%) | 75 (33.9%) | 438 (47.0%) | <0.01 |
|  | TNBC | 376 (31.4%) | 27 (58.7%) | 83 (37.6%) | 266 (28.5%) |  |
|  | HER2 | 295 (24.6%) | 4 (8.7%) | 63 (28.5%) | 228 (24.5%) |  |
| str TILs (mean) |  | 20.0 [10.0-30.0] | 20.0 [13.8-40.0] | 20.0 [10.0-40.0] | 15.0 [10.0-30.0] | 0.02 |
| IT TILs (mean) |  | 5.0 [5.0-15.0] | 5.0 [5.0-11.2] | 7.5 [5.0-20.0] | 5.0 [3.0-15.0] | 0.47 |
| NAC Regimen | AC | 235 (19.6%) | 4 (8.7%) | 25 (11.4%) | 206 (22.2%) | <0.01 |
|  | AC-Taxanes | 845 (70.7%) | 41 (89.1%) | 180 (81.8%) | 624 (67.1%) |  |
|  | Taxanes | 25 (2.1%) | 1 (2.2%) | 6 (2.7%) | 18 (1.9%) |  |
|  | Others | 91 (7.6%) | 0 (0.0%) | 9 (4.1%) | 82 (8.8%) |  |
| pCR class | No pCR | 911 (76.2) | 25 (54.3) | 157 (71.4) | 729 (78.4) | <0.001 |
|  | pCR | 285 (23.8) | 21 (45.7) | 63 (28.6) | 201 (21.6) |  |
| Nodal involvement | 0 | 682 (57.0) | 35 (76.1) | 141 (64.1) | 506 (54.4) | 0.003 |
|  | 1-3 | 341 (28.5) | 6 (13.0) | 58 (26.4) | 277 (29.8) |  |
|  | ≥4 | 174 (14.5) | 5 (10.9) | 21 (9.5) | 148 (15.9) |  |
| str TILs (mean) |  | 10.0 [5.0-15.0] | 15.0 [5.0-20.0] | 10.0 [5.0-15.0] | 10.0 [5.0-15.0] | 0.36 |
| IT TILs (mean) |  | 5.0 [2.0-10.0] | 5.0 [4.3-10.0] | 5.0 [2.0-10.0] | 5.0 [2.0-10.0] | 0.57 |

Missing data : Menopausal status, n=10; BMI (continuous), n=6; BMI class, n=6; Family history, n=3; Clinical tumor stage, n=1; Clinical nodal status, n=1; Histology, n=19; Grade, n=42; Mitotic index, n=502; Pre-NAC str TILs, n=482; Pre-NAC IT TILs, n=482; NAC regimen, n=3; pCR status, n=3; Post-NAC Nodal involvement, n=2; Post-NAC str TILs, n=482; Post-NAC IT TILs, n=714.
Abbreviations: NAC=neoadjuvant chemotherapy ; BMI=body mass index; NST= no special type; TNBC= triple negative breast cancer ; str TILs= stromal tumor-infiltrating lymphocytes ; IT TILs= intratumoral-infiltrating lymphocytes; AC=anthracyclines; pCR=Pathologic complete response.
The "n" denotes the number of patients. In case of categorical variables, percentages are expressed between brackets. In case of continuous variables, mean value is reported. In case of nonnormal continuous variables, median value is reported, with interquartile range between brackets.

Among the 267 screened patients, the distribution of *BRCA* status was as follows: *BRCA-*proficient n=221 (83%); *BRCA-*deficient, n= 46 (17%) (*BRCA1*-deficient, n=31 (67.39%); *BRCA2-*deficient, n = 14 (30.43%) and *BRCA1+2-*deficient, n=1 (2.17%)). Median age at diagnosis for patient with available *BRCA* mutation status was 40 years old (range 24-70) and most patients (n=227, 85%) were premenopausal. Patients repartition by subtype was as follows: luminal (n=90, 33.7%), TNBC (n= 110, 41.2%), *HER2*-positive (n= 67, 25.1%) (**Supplementary Table S1, Supplementary Fig. S2**).

Carriers of a *BRCA* pathogenic variant were more likely to have familial history of breast cancer (73.9% vs. 52.3%, p= 0.012), and to be diagnosed with TNBC (58.7% vs 37.6%; p= 0.006) than *BRCA*-proficient patients (**Table 1**). No other pattern among age, body mass index, histology, tumor size, nor proliferation indices (grade, mitotic index, KI67) was significantly different according to *BRCA* variant status. These results were substantially similar after the subgroup analysis of BC subtype (**Supplementary Table S2**).

**Table 2.**
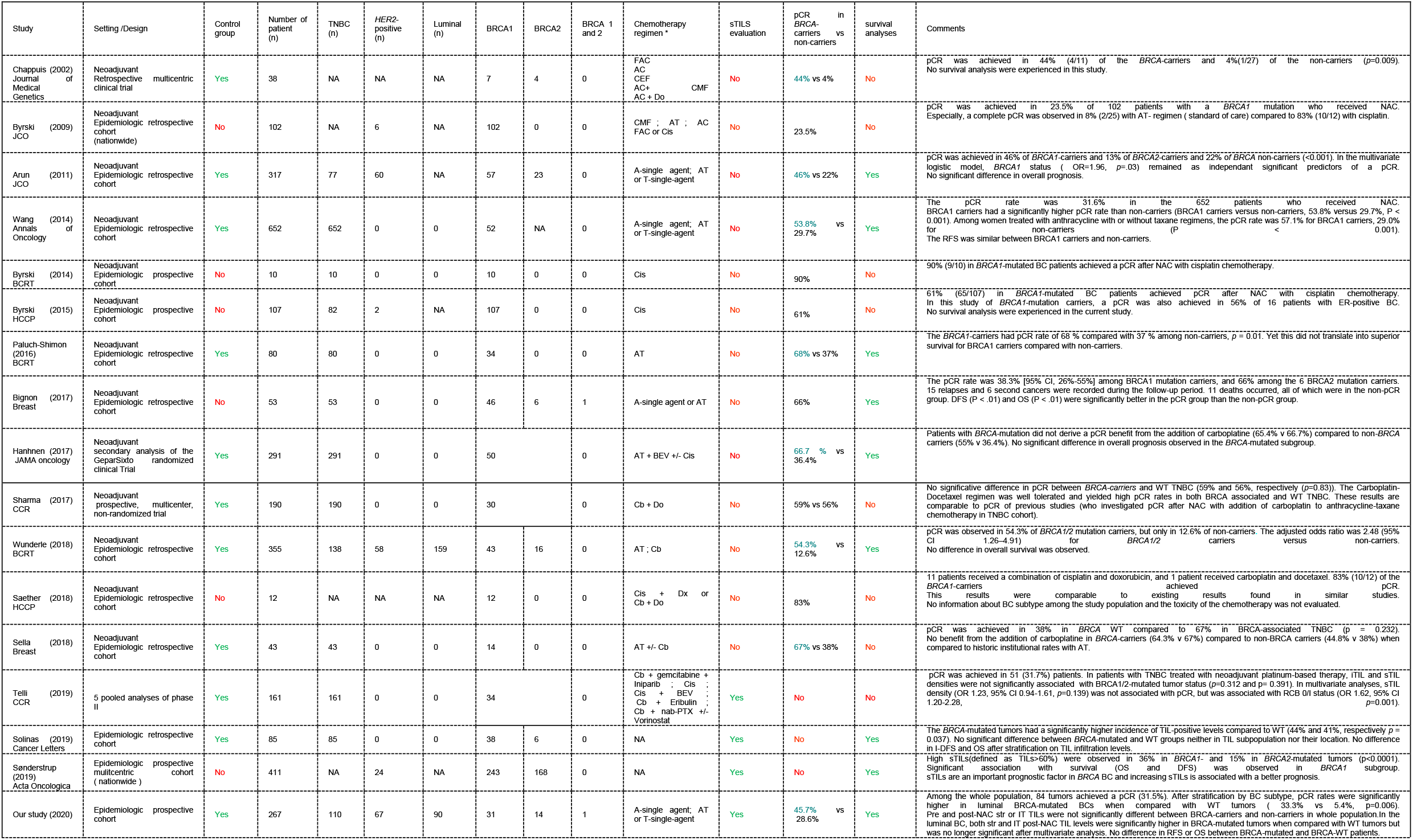
Literature Review. Abbreviations, CMF=cisplatine-methotrexate-fluorouracile; AT=docetaxel-doxorubicine; AC=doxorubicine-cyclophosphamide; FAC=fluorouracile-doxorubicin-cyclophophosphamide; CEF=cyclophosphamide-epirubicine-fluorouracile; A=anthracycline; Dx=doxorubicine; Do=docetaxel; Cb=carboplatin; Cis=cisplatine; BEV=bevacizumab; PTX=paclitaxel; T=taxane.

Baseline TILs were available for 192 out of 267 screened patients (72%). Neither pre-NAC str TIL levels (**Figs 1A-D**), nor IT TILs (**Figs 1E-H**) were significantly different by BRCA status (**Supplementary Table S1**), nor in each BC subtype (**Supplementary Table S2**). There was a strong, positive, linear relationship between stromal and intra-tumoral TILs (Spearman correlation coefficient of 0.74, *p<* 0.001, **Supplementary Fig. S3**)

**Figure 1.**
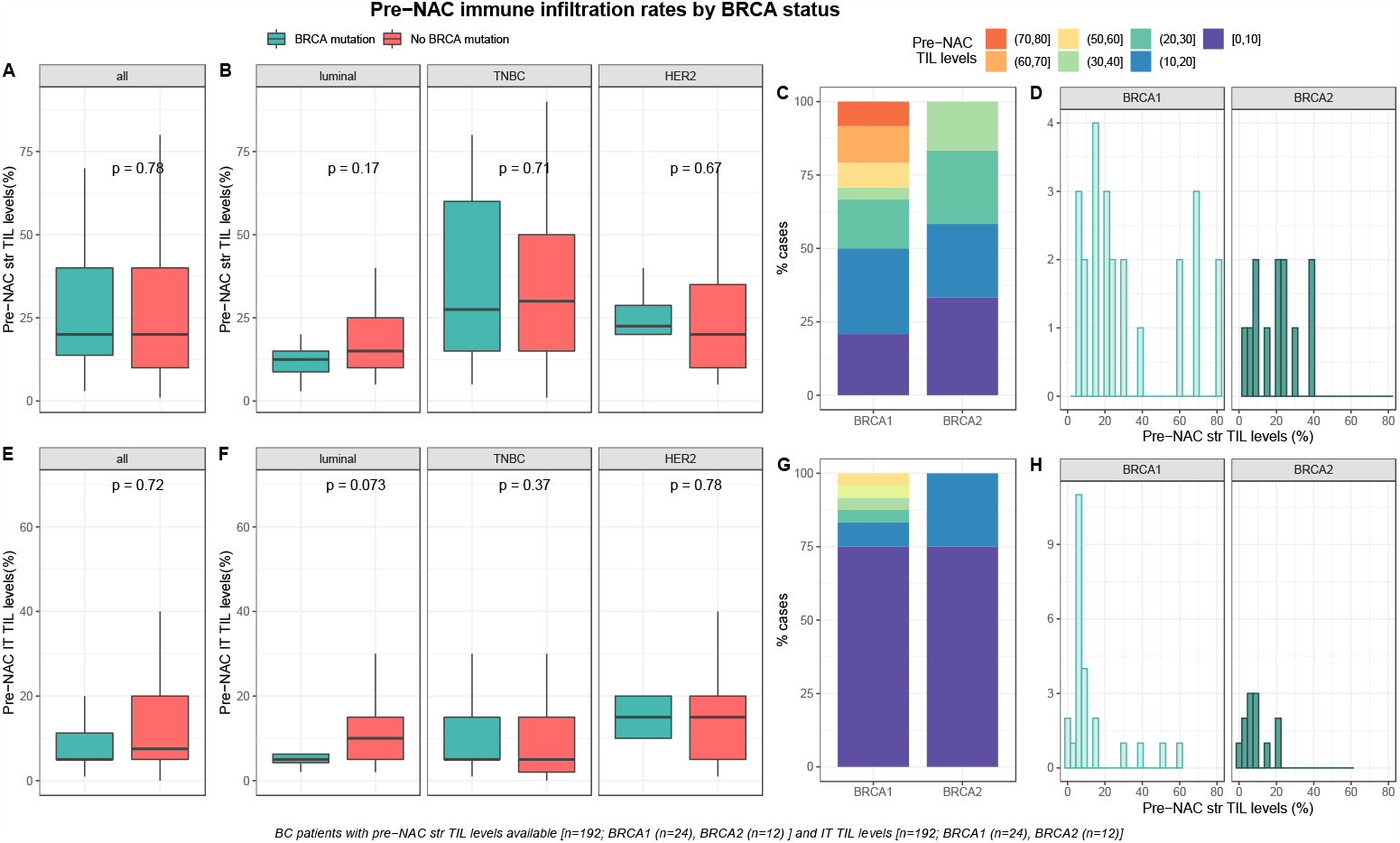
Associations between pre-NAC TILs and *BRCA* status in whole population, and by breast cancer subtype. Bottom and top bars of the boxplots represent the first and third quartiles, respectively, the medium bar is the median, and whiskers extend to 1.5 times the interquartile range. **A**, stromal lymphocytes among the whole population (All(n=192), *BRCA* mutation (n=36), *BRCA* wild-type(n=156). **B**, stromal lymphocytes in each BC subtype (Luminal(n=52), *BRCA* mutation(n=8), *BRCA* wild-type(n=44); TNBC(n=97), *BRCA* mutation(n=24), *BRCA* wild-type(n=73); *HER2*(n=43),*BRCA* mutation(n=4), *BRCA* wild-type(n=39). **C**, percentage of tumor according to pre-NAC stromal lymphocytes levels binned by 10% increment in patients with *BRCA*-deficient (*BRCA*1 (n=24), *BRCA*2(n=12)). **D**, distribution of pre-NAC stromal lymphocytes by gene mutations (histogram plot) in patients with *BRCA*-deficient (*BRCA*1 (n=24), *BRCA*2(n=12)). **E**, intratumoral lymphocytes among the whole population (All(n=192), *BRCA* mutation (n=36), *BRCA* wild-type(n=156)). **F**, intratumoral lymphocytes in each BC subtype (Luminal(n=52), *BRCA* mutation(n=8), *BRCA* wild-type(n=44); TNBC(n=97), *BRCA* mutation(n=24), *BRCA* wild-type(n=73); *HER2*(n=43),*BRCA* mutation(n=4), *BRCA* wild-type(n=39)). **G**, Percentage of tumor according to pre-NAC intratumoral lymphocytes levels binned by 10% increment in patients with *BRCA*-deficient (*BRCA*1 (n=24), *BRCA*2(n=12)). **H**, distribution of pre-NAC intratumoral lymphocytes by gene mutations (histogram plot) in patients with *BRCA*-deficient (*BRCA*1 (n=24), *BRCA*2(n=12)).

### Response to treatment and post-NAC immune infiltration

#### Response to treatment

At NAC completion, pCR was observed in 84 out of 266 (31%) patients and pCR rates were significantly different by BC subtype (luminal: 10% (9/89), TNBC: 45% (49/110) and *HER2-* positive 39% (26/67), *p<* 0.001). Pre-NAC str TIL levels were significantly higher in tumors for which pCR was achieved (*p<* 0.001) and there was a significant association between pre-NAC TIL levels and pCR status in the whole population (all: OR = 1.03, CI95% [1.02 – 1.05], *p<* 0.001; Supplementary Fig. S3) and in the TNBC subgroup (luminal: OR = 1.03, CI95% [1 – 1.09], *p=* 0.21; TNBC: OR = 1.03; CI95% [1–1.04], *p=* 0.007; *HER2-*positive: OR = 1.02, CI95% [0.99–1.06], *p=* 0.23; **Supplementary Fig. S4**).

pCR rates were significantly higher in patients with *BRCA-*deficient breast cancers (45.7% (21/46) versus 28 % (63/221) in *BRCA-*proficient, *p<* 0.035, **Supplementary Table S1, Figure 2**). After the subgroup analysis of BC subtype, this was confirmed only in the luminal BC subtype (33.3% (5/15), *p=* 0.006), but not in TNBC and *HER2*-positive BCs (48.1% (13/27), *p=* 0.823 and 75% (3/4), *p=* 0.291, respectively, **Supplementary Table S2, Figure 2**). The interaction test between BC subtype and *BRCA* status was nearly significant (*P*_*interaction*_=0.056).

**Figure 2.**
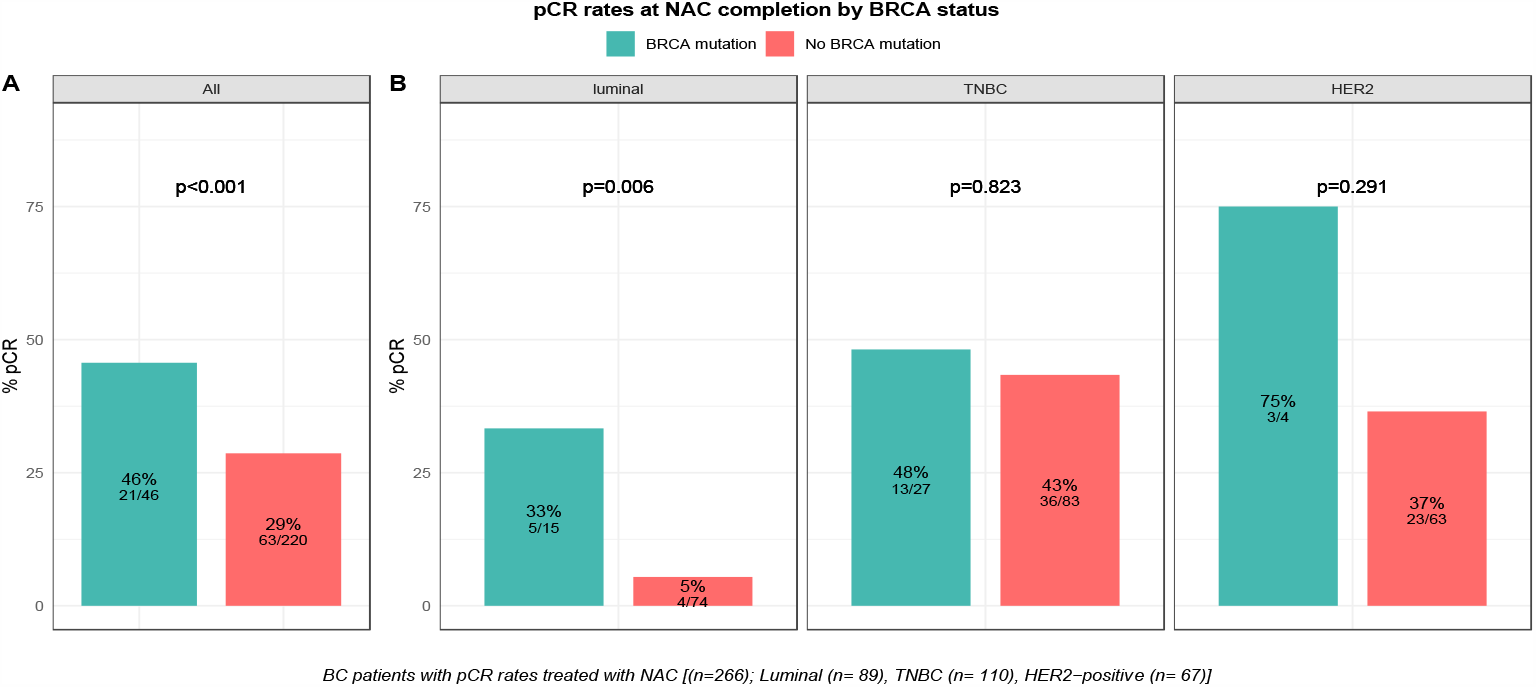
Barplot of associations between response to treatment and *BRCA* status in whole population, and by breast cancer subtype. **A**, among the whole population (All(n=266), *BRCA* mutation (n=46), *BRCA* wild-type(n=220)). **B**, by BC subtype (Luminal(n=89), *BRCA* mutation(n=15), *BRCA* wild-type(n=74); TNBC(n=110), *BRCA* mutation(n=27), *BRCA* wild-type(n=83); *HER2*(n=67),*BRCA* mutation(n=4), *BRCA* wild-type(n=63)).

However, *BRCA* status was not significantly associated with pCR after multivariate analysis, and only BC subtype (TNBC, OR = 7.14, CI95% [3.39 - 16.57], *p<* 0.001; *HER2*-positive, OR = 5.64, CI95% [2.5 - 13.78], *=*<0.001), tumor size (T2, OR = 0.37, CI95% [0.16 - 0.83], *p=* 0.017; T3, OR = 0.21, CI95% [0.08 - 0.55], *p=* 0.002) and pre-NAC str and IT TILs (OR = 1.03, CI95% [1.02 - 1.05], *p=* 0.001 and OR = 1.04, CI95% [1.02 - 1.07], *p=* 0.002) were independent predictors of pCR (**Supplementary Table S3**).

#### Post-NAC Immune Infiltration by BRCA status

After NAC, str and IT TILs were available in 192 (72%) and 120 (45%) patients respectively. Post-NAC immune infiltration (whether intra-tumoral or stromal) was not significantly different between *BRCA-*deficient and *BRCA-*proficient carriers (**Supplementary Table S1, Fig. 3A-3E**). However, both str and IT TIL levels were significantly higher in tumors with *BRCA* pathogenic mutations when compared with wild-type tumors in luminal BCs (median str TIL levels: 15% vs. 10%, *p=* 0.009 and median IT TIL levels: 10% vs. 5%, *p=* 0.019, respectively, **Supplementary Table S2, Figure 3**).

**Figure 3.**
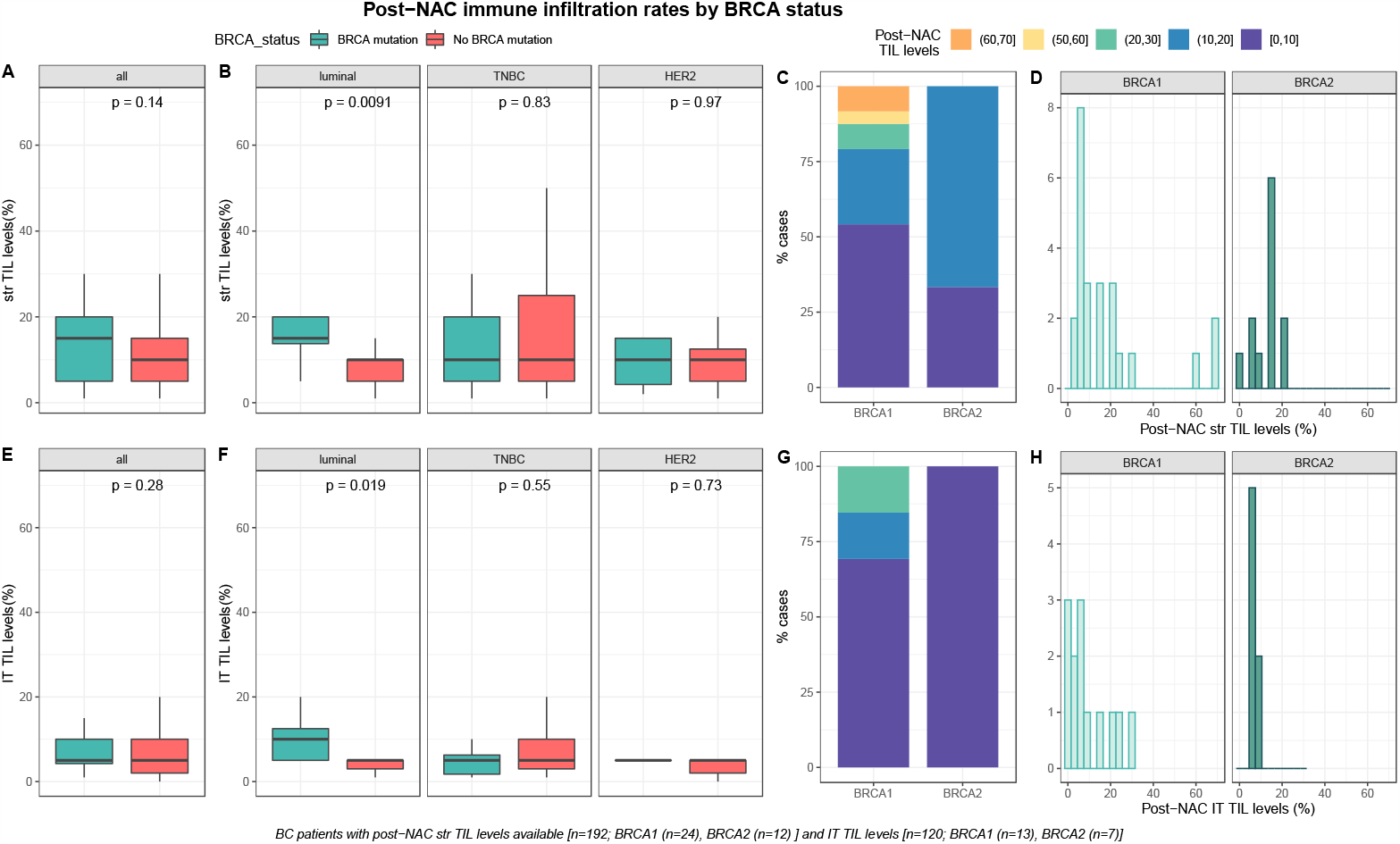
Associations between post-NAC TILs and *BRCA* status in whole population, and after stratification by breast cancer subtype. Bottom and top bars of the boxplots represent the first and third quartiles, respectively, the medium bar is the median, and whiskers extend to 1.5 times the interquartile range. **A**, stromal lymphocytes among the whole population (All(n=192), *BRCA* mutation (n=36), *BRCA* wild-type(n=156)). **B**, stromal lymphocytes in each BC subtype (Luminal(n=52), *BRCA* mutation(n=8), *BRCA* wild-type(n=44); TNBC(n=97), *BRCA* mutation(n=24), *BRCA* wild-type(n=73); *HER2*(n=43),*BRCA* mutation(n=4), *BRCA* wild-type(n=39)). **C**, Percentage of tumor according to post-NAC stromal lymphocytes levels binned by 10% increment in patients with B*RCA*-deficient (*BRCA*1 (n=24), *BRCA*2(n=12)). **D**, distribution of post-NAC stromal lymphocytes by gene mutations (histogram plot) in patients with *BRCA*-deficient (*BRCA*1(n=24), *BRCA*2(n=12)). **E**, intratumoral lymphocytes among the whole population (All(n=120), *BRCA* mutation (n=20), *BRCA* wild type(n=100)). **F**, intratumoral lymphocytes in each BC subtype (Luminal(n=44), *BRCA* mutation(n=7), *BRCA* wild-type(n=37); TNBC(n=50), *BRCA* mutation(n=12), *BRCA* wild-type(n=38); *HER2*(n=26),*BRCA* mutation(n=1), *BRCA* wild-type(n=25)). **G**, percentage of tumor according to post-NAC intratumoral lymphocytes levels binned by 10% increment in patients with *BRCA*-deficient (*BRCA*1 (n=13), *BRCA*2(n=7)). **H**, distribution of pre-NAC intratumoral lymphocytes by gene mutations (histogram plot) in patients with *BRCA*-deficient (*BRCA*1 (n=13), *BRCA*2(n=7)).

Median pre-NAC str TIL were higher than after NAC (20% vs 10%, 11.95%), also according to *BRCA* status and type (**Supplementary Table S1, Fig. 4**). There was no correlation between pre and post NAC str TILs (correlation coefficient of 0.13 and *p<* 0.06, **Supplementary Fig. S5A**) and there was a weak, positive, linear relationship between pre and post NAC IT TIL levels (correlation coefficient of 0.31 and *p<* 0.001, **Supplementary Fig. S5B**).

**Figure 4.**
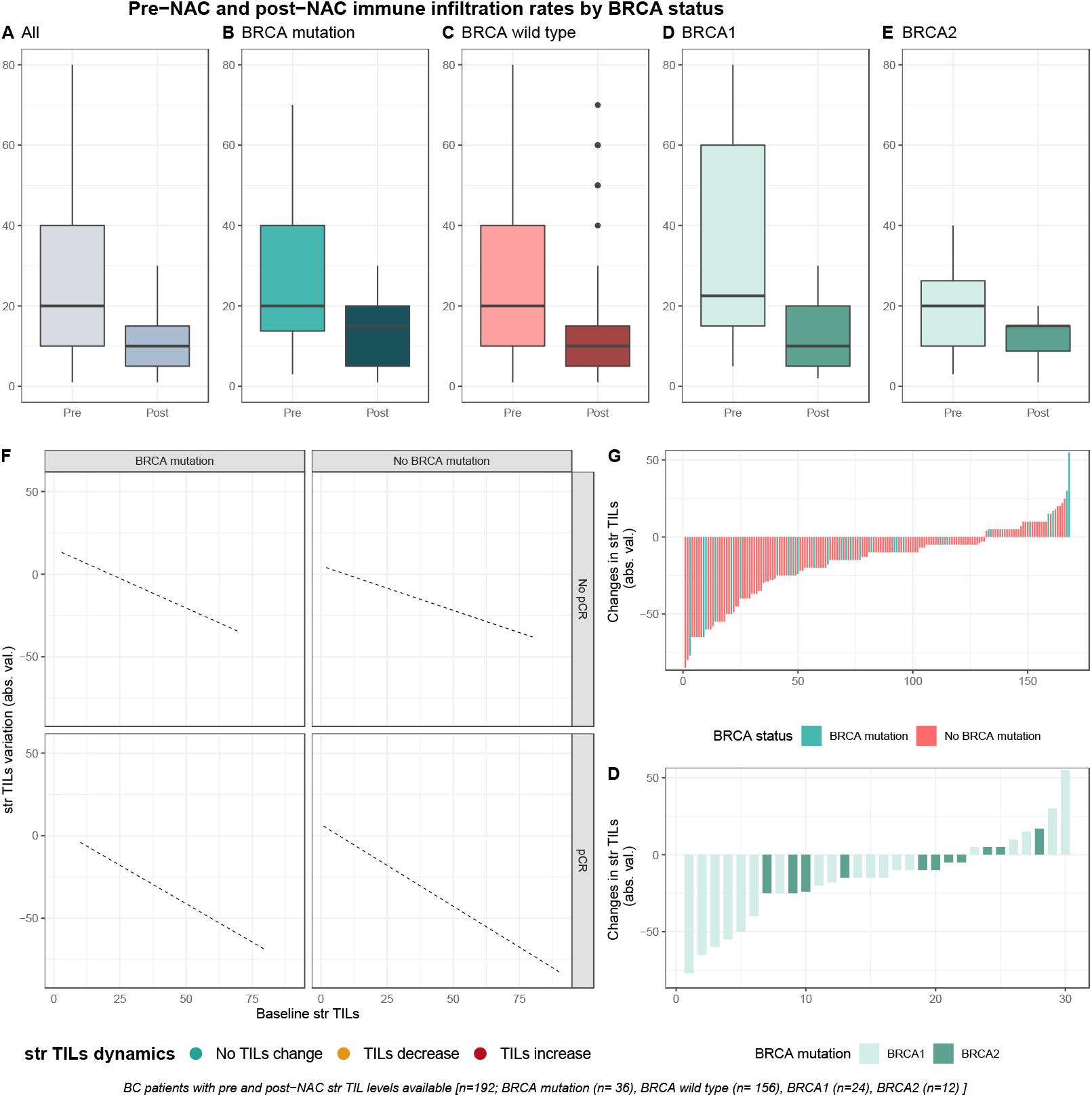
Pre-NAC and post-NAC stromal immune infiltration rates in the whole population and by BRCA status. **A-E**, bar plots of str TIL levels before and after NAC in the whole population and in *BRCA* pathogenic variant. Bottom and top bars of the boxplots represent the first and third quartiles, respectively, the medium bar is the median, and whiskers extend to 1.5 times the interquartile range. (All(n=192); *BRCA* mutation (n=36), *BRCA* wild-type(n=156); *BRCA*1(n=24), *BRCA*2(12)). **F**, variation of str TIL levels according to the pre-NAC str TIL levels binned by BRCA status and response to chemotherapy. Points represent the difference between pre- and post-NAC paired TIL levels values of a given patient and are colored according to TIL variation category (TIL level decrease: yellow/no change: green/increase: red) (All(n=191), *BRCA* mutation (n=36), *BRCA* wild-type(n=155)). **G-D**, waterfall plot representing the variation of TIL levels according to *BRCA*-deficient (*BRCA*1-deficient, *BRCA*2-deficient); each bar represents one sample, and samples are ranked by increasing order of TIL level change. Paired samples for which no change was observed have been removed from the graph. (All(n=191), *BRCA* mutation [(n=36), *BRCA*1, n= 24; *BRCA*2= 12)], *BRCA* wild-type(n=155)).

### Survival analysis

After a median of follow-up of 90.4 months (range from 0.2 to 187 months), 73 patients experienced relapse, and 38 died. RFS and OS were not significantly different between carriers of a *BRCA* pathogenic variant and *BRCA*-proficient patients, neither were they in screened population nor after the subgroup analysis of BC subtype (**Supplementary Figs. S6-7)**.

## Discussion

In the current study, we did not identify any association between *BRCA* status and immune infiltration whatever the type of TILs (IT, str). We found a better response to neoadjuvant chemotherapy in tumors associated with a germline *BRCA* pathogenic variant when compared to *BRCA*-WT, but the latter was limited to the group of luminal BCs and was not statistically significant after multivariate analysis. Probably in relation, we recovered higher post-NAC lymphocyte infiltration in *BRCA*-deficient tumors in the luminal BC subgroup.

Regarding pre-treatment immune infiltration, Sønderstrup and colleagues (25) analyzed str TIL levels in a nationwide cohort of *BRCA1* and *BRCA2* carriers with primary BCs. They found a greater prevalence of high stromal TILs (defined as TILs-positive tumors with ≥ 60% str TILs) in *BRCA1*-deficient tumors (n=243) when compared with *BRCA2*-deficient tumors (n=168) (36% *versus* 15 % respectively, *p* <0.0001). However, no control group with *BRCA*-WT tumors was available in this study. In a small study of 85 TNBC patients, Solinas and colleagues (26) investigated the distribution of TILs subpopulations. The tumors of patients in the *BRCA1* or *BRCA2*-mutated group displayed a higher prevalence of TILs-positive tumors (defined as tumors with ≥ 10% str or IT TILs) when compared with the *BRCA*-WT (93.2% *versus* 75.6% respectively, *p*=0.037). No other statistically significant differences were identified between *BRCA*-carriers and non-carriers, neither in TILs subpopulations nor their location. More recently, Telli and colleagues (27) investigated the association between TILs, homologous recombination deficiency (HDR) and *BRCA1*/2 status in a cohort of 161 TNBC patients pooled from 5 phase II neoadjuvant clinical trials of platinum-based therapy. They found that IT TILs and str TILs density were not associated with BRCA1/2 status (*p=*0.312 and *p=* 0.391, respectively). Consistently with Telli *et al*, we did not observe any difference in baseline immune infiltration according to *BRCA* status.

Some retrospective studies suggested that tumors displayed higher chemosensitivity according to *BRCA*-mutation status (17–19, 28–35). Arun *et al*. (31) compared pCR rates after NAC between *BRCA1* or *BRCA2*-carriers (n=57 and n=23, respectively) and WT controls (n=237). The majority of patients (82%) received an anthracycline-taxane containing regimen as NAC. The authors found that *BRCA1* mutation was an independent positive predictor of pCR (OR=3.16, 95%CI 1.55-6.42, *p=* 0.002). In the largest study so far, Wunderle *et al*.(18) investigated efficacy of chemotherapy among a cohort of 355 patients composed with 16.6% (59/355) of *BRCA*-carriers. Across all BC subtypes, 64.4% of patients with a *BRCA1/2* pathogenic variant received anthracycline-based treatments, while the rest received carboplatin. pCR was observed in 54.3% (32/59) of all *BRCA1/2* mutation carriers, and in 39.5% (15/34) of the *BRCA*-carriers *versus* 13% of the WT BCs in the anthracycline-regimen. In our cohort, we found similar results after univariate analysis, and we additionally evidenced a nearly significant interaction with BC subtype. The fact that our results were no longer significant after multivariate analysis is possibly due to a lack of statistical power.

Furthermore, we found that both str and IT TIL levels were higher after NAC completion in the luminal BCs. Whether this difference in post treatment TILs is a cause, a consequence, or unrelated to response to chemotherapy remains unknown. Indeed, post-NAC TIL levels have been shown to be strongly related to response to chemotherapy in BC cohorts including all BC subtypes (36–38) but only a few studies have investigated the dynamic of TIL levels in response to NAC. Hamy *et al*.(38) noticed that mean TIL levels decreased after chemotherapy completion across all the BC subtype (pre-NAC TILs: 24.1% vs. post-NAC TILs: 13.0%, *p<* 0.001). This decrease was strongly associated with high pCR rates, and the variation of TIL levels was strongly inversely correlated with pre-NAC TIL levels (and the variation of TIL levels was strongly inversely correlated with pre-NAC TIL levels (r= − 0.80, *p<* 0.001).

Finally, in line with several recently published clinical studies (39–41), we found that survival outcomes were not different between *BRCA*-carriers and non-carriers. A multivariate study, including 223 BC patients carrying *BRCA* pathogenic variants and 446 controls with sporadic BC matched for age and year of diagnosis, showed no difference in terms of specific BC survival between *BRCA1* or *BRCA2* mutation carriers and controls (42). Templeton et *al*. evaluated a total of 16 studies comprising data from 10,180 patients and concluded that *BRCA* pathogenic mutations were not associated with a worse overall survival (43).

Limits of our study include its retrospective design as well as small effectives potentially leading to a lack of statistical power. Moreover the incidence of bi-allelic pathogenic alterations in HR-related genes according to somatic origin is well-known and ranches from 1 to 2 % (44) but we did not explore somatic mutational status in the tumor tissues in the current study.

It also has several strengths, for instance by being the largest cohort with a *BRCA*-WT control group, and analyses performed after stratification by BC subtype. Finally, to our knowledge, we provide data on post-NAC immune infiltration according to *BRCA* status for the first time.

Our study has several implications. First, it generates an unprecedented hypothesis that luminal BC patients with germline *BRCA* pathogenic variants may represent a subset of luminal BCs that are more likely to benefit from chemotherapy as primary treatment than the whole luminal BC population. It is known that the absolute benefit of chemotherapy is lower in luminal BC than in the other BC subtypes (45). If further validated in independent cohorts, our findings might lead to reconsider standard use of chemotherapy in patients with luminal BC associated with *BRCA* pathogenic mutations. Second, patients not achieving pCR may be candidates for post-operative clinical trials exploring alternative therapeutic strategies. As post-NAC immune infiltration seems to be higher in post-NAC specimens of luminal tumors with *BRCA* pathogenic mutations, we can hypothesize that those tumors would be more likely to respond to checkpoint inhibitors after chemotherapy. Second line trials using immune checkpoint inhibitors (such as anti–PD-1 and anti–PD-L1 antibodies) alone or in combination, together with endocrine therapy could be a relevant strategy for patients failing to reach pCR at NAC completion.

## Data Availability

All data are available in open recquest.

## Supplementary material

### 1. Patients and treatments

#### 1.1. Patients

In total, patients with T1-3NxM0 invasive breast cancer (BC) (NEOREP Cohort, CNIL declaration number 1547270) treated at Institut Curie (Paris and Saint Cloud) between 2002 and 2012 were included in this study. We included unilateral, non-recurrent, non-inflammatory, non-metastatic tumors, excluding T4 tumors. NAC regimens changed over time (anthracycline-based regimen or sequential anthracycline-taxane regimen) with trastuzumab used in an adjuvant and/or neoadjuvant setting since 2005 for *HER2-*positive tumors. All patients underwent radiotherapy. Endocrine therapy (tamoxifen or aromatase inhibitor) was prescribed when indicated. This study was approved by the Breast Cancer Study Group of Institut Curie.

#### 1.2. Treatments

NAC regimens changed over time (anthracycline-based regimen or sequential anthracycline-taxane regimen), with trastuzumab used in an adjuvant and/or neoadjuvant setting for HER2-positive tumors since the middle of the past decade. Trastuzumab treatments changer over time due to a change of marketing authorization during the study period. Adjuvant hormone therapy (tamoxifen, aromatase inhibitor, or GnRH agonist) was prescribed when indicated. Surgery (breast-conserving or mastectomy) was performed 4-6 weeks after NAC. Every patient received adjuvant radiotherapy. Adjuvant chemotherapy (ADJ) was decided after multidisciplinary consultation meeting considering patient characteristic, prognosis factor and response to NAC (residual disease and/or node involvement). Patient follow-up after treatment was of every 4 months during the first 2 years, then every 6 months for 3 years, and once a year starting from the 5^th^ year. Follow-up consisted of clinical examination associated to mammography and mammary ultrasound once a year, with annual Magnetic resonance imaging (RMI) in *BRCA*-carriers.

### 2. Tumor samples and pathological review

#### 2.1. ER, PR, HER2 status and BC subtype

Cases were considered to be estrogen receptor (ER)-positive or progesterone receptor (PR)-positive if at least 10% of the tumor cells expressed estrogen and/or progesterone receptors (ER/PR). *HER2* expression was determined by immunohistochemistry, with scoring according to the American Society of Clinical Oncology (ASCO)/College of American Pathologists (CAP) guidelines (1). Scores of 3+ were reported as positive, and scores of 1+/0 as negative. Tumors with scores of 2+ were further tested by fluorescence *in situ* hybridization (FISH). For *HER2* gene amplification, we evaluated a mean of 40 tumor cells per sample and calculated the mean *HER2* signal per nucleus. A *HER2*/CEN17 ratio ≥ 2 was considered positive, and a ratio < 2 was considered negative (1).

#### 2.2. Other pathological parameters

Histological grade was determined as described by Elston Ellis. Mitotic cells were counted on 10 high-power fields (HPF) (x40 objective; field diameter = 0.62 mm) and cutoffs of <11, 12– 22 and >22 mitoses were used to define low, intermediate and high mitotic indices, respectively, according to the international recommendations(2). Due to significant differences in distribution before and after NAC, invasive tumor cellularity was binned according to the median value (pre-NAC: 60%; post-NAC: 30%).

#### 2.3 BRCA status

Since 2002, patient referral for genetic counseling depends on individual or family criteria. These criteria are based on the probability of identifying a genetic predisposition in the family of at least 10% (in particular a germline *BRCA1* or *BRCA 2* pathogenic variant). The individual criteria are: early age at diagnosis (under 40) or bilateral breast cancer: synchronous or metachronous (with the first breast cancer before age 50), or specific phenotype (triple negative cancer before age 51).The family criteria are: 3 cases of breast cancer in the same branch of heredity, or 2 cases of breast cancer including 1 under 45-50, of breast or ovarian cancer, or 2 cases of breast cancer including 1 male. The 2 cases are women relatives of the first degree (or second degree if paternal transmission).

#### 2.4 TILs levels

Infiltrates were scored on a continuous scale, as the mean percentage of the stromal area occupied by mononuclear cells. After NAC, we assessed TIL levels within the borders of the residual tumor bed, as defined by the RCB index(3). Nothing is known about the clinical, biological and prognostic significance of TILs in the area of regression in cases of pathological response, but the TILs international working group recently called for their evaluation for research purposes. In cases of pCR, the scar area was measured on macroscopic examination. The scar appeared as a white area in the breast parenchyma corresponding to the tumor bed modified by NAC. It was characterized by the presence of histiocytes, lymphocytes, macrophages, fibrosis and elastosis. The whole fibro-inflammatory scar was evaluated on HE sections (size in mm and stromal TIL level evaluation).

## Supplementary Tables

**Supplementary Table S1.** BRCA screened patients’characteristics among by BRCA status

| Characteristics | Class | Overall | BRCA mutation | BRCA wild-type | p |
| --- | --- | --- | --- | --- | --- |
| n= |  | 267 (100%) | 46(17%) | 221(83%) |  |
| Age (mean) |  | 41.31 | 39.5 | 41.7 | 0.15 |
| Menopausal status | pre | 228 (85.7%) | 41 (89.1%) | 187 (85.0%) | 0.62 |
|  | post | 38 (14.3%) | 5 (10.9%) | 33 (15.0%) |  |
| BMI (mean) |  | 23.50 | 22.8 | 23.6 | 0.24 |
| BMI class | [15,19] | 23 (8.6%) | 6 (13.3%) | 17 (7.7%) | 0.248 |
|  | (19,25] | 178 (66.9%) | 31 (68.9%) | 147 (66.5%) |  |
|  | (25,30] | 47 (17.7%) | 4 (8.9%) | 43 (19.5%) |  |
|  | (30,50] | 18 (6.8%) | 4 (8.9%) | 14 (6.3%) |  |
| Family history of BC | no | 116 (43.9%) | 12 (26.1%) | 104 (47.7%) | 0.01 |
|  | yes | 148 (56.1%) | 34 (73.9%) | 114 (52.3%) |  |
| Clinical tumor size | T1 | 27 (10.1%) | 5 (10.9%) | 22 (10.0%) | 0.50 |
|  | T2 | 181 (67.8%) | 28 (60.9%) | 153 (69.2%) |  |
|  | T3 | 59 (22.1%) | 13 (28.3%) | 46 (20.8%) |  |
| Clinical nodal status | N0 | 110 (41.2%) | 17 (37.0%) | 93 (42.1%) | 0.63 |
|  | N1-N2-N3 | 157 (58.8%) | 29 (63.0 %) | 128 (57.9%) |  |
| Histology | NST | 256 (95.9%) | 43 (93.5%) | 213 (96.4%) | NaN |
|  | others | 11 (4.1%) | 3 (6.5%) | 8 (3.6%) |  |
| Grade | I-II | 86 (32.8%) | 10 (23.3%) | 76 (34.7%) | 0.24 |
|  | III | 176 (67.2%) | 33 (76.7%) | 143 (65.3%) |  |
| Mitotic Index (mean) |  | 26.57 | 30.8 | 25.6 | 0.24 |
| Subtype | luminal | 90 (33.7%) | 15 (32.6%) | 75 (33.9%) | <0.01 |
|  | TNBC | 110 (41.2%) | 27 (58.7%) | 83 (37.6%) |  |
|  | HER2 | 67 (25.1%) | 4 (8.7%) | 63 (28.5%) |  |
| str TILs (mean) |  | 20.0 [10.0-40.0] | 20.0 [13.8-40.0] | 20.0 [10.0-40.0] | 0,78 |
| IT TILs (mean) |  | 5.0 [5.0-15.0] | 5.0 [5.0-11.3] | 7.5 [5.0-20.0] | 0,72 |
| NAC Regimen | AC | 29 (10.9%) | 4 (8.7%) | 25 (11.4%) | 0.49 |
|  | AC-Taxanes | 221 (83.1%) | 41 (89.1%) | 180 (81.8%) |  |
|  | Taxanes | 7 (2.6%) | 1 (2.2%) | 6 (2.7%) |  |
|  | Others | 9 (3.4%) | 0 (0.0%) | 9 (4.1%) |  |
| pCR class | No pCR | 182 (68.4%) | 25 (54.3%) | 157 (71.4%) | 0,04 |
|  | pCR | 84 (31.6%) | 21 (45.7%) | 63 (28.6%) |  |
| Nodal involment | 0 | 176 (66.2%) | 35 (76.1%) | 141 (64.1%) | 0,16 |
|  | 1-3 | 64 (24.1%) | 6 (13.0%) | 58 (26.4%) |  |
|  | ≥4 | 26 (9.8%) | 5 (10.9%) | 21 (9.5%) |  |
| str TILs (mean) |  | 10.0 [5.0-15.0] | 15.0 [5.0-20.0] | 10.0 [5.0-15.0] | 0,14 |
| IT TILs (mean) |  | 5.0 [2.0-10.0] | 5.0 [4.3-10.0] | 5.0 [2.0-10.0] | 0,27 |

Missing data: Menopausal status, n=1; BMI (continuous), n=1; BMI class, n=1; Family history, n=3; Grade, n=5; Mitotic index, n=77; Pre-NAC str TILs, n=75; Pre-NAC IT TILs, n=75; NAC regimen, n=1; pCR status, n=1; Post-NAC Nodal involment, n=1; Post-NAC str TILs, n=75; Post-NAC IT TILs, n=147.
NAC=neoadjuvant chemotherapy ; BMI=body mass index; NST=no special type ; TNBC= triple negative breast cancer ; str TILs= stromal tumor-infiltrating lymphocytes ; IT TILs= intratumoral-infiltrating lymphocytes; AC=anthracyclines; pCR=Pathologic complete response.
The “n” denotes the number of patients. In case of categorical variables, percentages are expressed between brackets. In case of continuous variables, mean value is reported. In case of nonnormal continuous variables, median value is reported, with interquartile range between brackets.

**Supplementary Table S2.** Patients’characteristics in each tumor subtype and by BRCA status

| Characteristics | Class | Luminal |  |  | TNBC |  |  | HER2 |  |  |
| --- | --- | --- | --- | --- | --- | --- | --- | --- | --- | --- |
|  |  | BRCA mutation | BRCA wild-type | p | BRCA mutation | BRCA wild-type | p | BRCA mutation | BRCA wild-type | p |
| n= |  | 15 (16.7%) | 75 (83.3%) |  | 27 (24.5%) | 83 (75.5%) |  | 4 (6.0%) | 63 (94.0%) |  |
| Age (mean) |  | 38.5 | 42.3 | 0,13 | 39.6 | 42.2 | 0,19 | 43.5 | 40.2 | 0,50 |
| Menopausal status | pre | 14 (93.3%) | 64 (86.5%) | 0,76 | 24 (88.9%) | 72 (86.7%) | 1,00 | 3 (75.0%) | 51 (81.0%) | 1,00 |
|  | post | 1 (6.7%) | 10 (13.5%) |  | 3 (11.1%) | 11 (13.3%) |  | 1 (25.0%) | 12 (19.0%) |  |
| BMI (mean) |  | 22.2 | 23.5 | 0,32 | 23.5 | 23.3 | 0,74 | 20.4 | 24.3 (4.4%) | 0,09 |
| BMI class | [15,19] | 4 (28.6%) | 7 (9.3%) | 0.204 | 1 (3.7%) | 7 (8.4%) | 0.251 | 1 (25.0%) | 3 (4.8%) | 0.298 |
|  | (19,25] | 8 (57.1%) | 49 (65.3%) |  | 20 (74.1%) | 55 (66.3%) |  | 3 (75.0%) | 43 (68.3%) |  |
|  | (25,30] | 1 (7.1%) | 14 (18.7%) |  | 3 (11.1%) | 18 (21.7%) |  | 0 (0.0%) | 11 (17.5%) |  |
|  | (30,50] | 1 (7.1%) | 5 (6.7%) |  | 3 (11.1%) | 3 (3.6%) |  | 0 (0.0%) | 6 (9.5%) |  |
| Family history | no | 2 (13.3%) | 26 (35.1%) | 0,18 | 10 (37.0%) | 46 (56.1%) | 0,13 | 0 (0.0%) | 32 (51.6%) | 0,14 |
|  | yes | 13 (86.7%) | 48 (64.9%) |  | 17 (63.0%) | 36 (43.9%) |  | 4 (100.0%) | 30 (48.4%) |  |
| Clinical tumor size | T1 | 1 (6.7%) | 5 (6.7%) | 0,99 | 2 (7.4%) | 11 (13.3%) | 0,28 | 2 (50.0%) | 6 (9.5%) | 0,04 |
|  | T2 | 11 (73.3%) | 54 (72.0%) |  | 16 (59.3%) | 56 (67.5%) |  | 1 (25.0%) | 43 (68.3%) |  |
|  | T3 | 3 (20.0%) | 16 (21.3%) |  | 9 (33.3%) | 16 (19.3%) |  | 1 (25.0%) | 14 (22.2%) |  |
| Clinical nodal status | N0 | 5 (33.3%) | 35 (46.7%) | 0,51 | 10 (37.0%) | 34 (41.0%) | 0,89 | 2 (50.0%) | 24 (38.1%) | 1,00 |
|  | N1-N2-N3 | 10 (66.7%) | 40 (53.3%) |  | 17 (63.0%) | 49 (59.0%) |  | 2 (50.0%) | 39 (61.9%) |  |
| Histology | NST | 13 (86.7%) | 72 (96.0%) | NaN | 26 (96.3%) | 78 (94.0%) | NaN | 4 (100.0%) | 63 (100.0%) | NaN |
|  | others | 2 (13.3%) | 3 (4.0%) |  | 1 (3.7%) | 5 (6.0%) |  | 0 (0.0%) | 0 (0.0%) |  |
| Grade | I-II | 6 (42.9%) | 42 (56.0%) | 0,55 | 2 (8.0%) | 10 (12.2%) | 0,53 | 2 (50.0%) | 24 (38.7%) | NaN |
|  | III | 8 (57.1%) | 33 (44.0%) |  | 23 (92.0%) | 72 (87.8%) |  | 2 (50.0%) | 38 (61.3%) |  |
| Mitotic Index (mean) |  | 23.0 | 18.5 | 0,56 | 36.3 | 31.8 | 0,46 | 13.0 | 21.5 | 0,45 |
| str TILs (mean) |  | 12.5 [8.8-15.0] | 15.0 [10.0-25.0] | 0,17 | 27.5 [15.0-60.0] | 30.0 [15.0-50.0] | 0,71 | 22.5 [20.0-28.8] | 20.0 [10.0-35.0] | 0,66 |
| IT TILs (mean) |  | 5.0 [4.3-6.3] | 10.0 [5.0-15.0] | 0,07 | 5.0 [5.0-15.0] | 5.0 [2.0-15.0] | 0,37 | 15.0 [10.0-20.0] | 15.0 [5.0-20.0] | 0,77 |
| NAC Regimen | AC | 3 (20.0%) | 13 (17.6%) | 0,83 | 1 (3.7%) | 9 (10.8%) | NaN | 0 (0.0%) | 3 (4.8%) | 0,77 |
|  | AC-Taxanes | 11 (73.3%) | 58 (78.4%) |  | 26 (96.3%) | 73 (88.0%) |  | 4 (100.0%) | 49 (77.8%) |  |
|  | Taxanes | 1 (6.7%) | 2 (2.7%) |  | 0 (0.0%) | 0 (0.0%) |  | 0 (0.0%) | 4 (6.3%) |  |
|  | Others | 0 (0.0%) | 1 (1.4%) |  | 0 (0.0%) | 1 (1.2%) |  | 0 (0.0%) | 7 (11.1%) |  |
| pCR class | No pCR | 10 (66.7%) | 70 (94.6%) | <0,01 | 14 (51.9%) | 47 (56.6%) | 0,83 | 1 (25.0%) | 40 (63.5%) | 0,32 |
|  | pCR | 5 (33.3%) | 4 (5.4%) |  | 13 (48.1%) | 36 (43.4%) |  | 3 (75.0%) | 23 (36.5%) |  |
| Nodal involvement | 0 | 8 (53.3%) | 30 (40.5%) | 0,23 | 23 (85.2%) | 65 (78.3%) | 0,74 | 4 (100.0%) | 46 (73.0%) | 0,49 |
|  | 1-3 | 3 (20.0%) | 32 (43.2%) |  | 3 (11.1%) | 13 (15.7%) |  | 0 (0.0%) | 13 (20.6%) |  |
|  | ≥4 | 4 (26.7%) | 12 (16.2%) |  | 1 (3.7%) | 5 (6.0%) |  | 0 (0.0%) | 4 (6.3%) |  |
| str TILs (mean) |  | 15.0 [13.8-20.0] | 10.0 [5.0-10.0] | <0,01 | 10.0 [5.0-20.0] | 10.0 [5.0-25.0] | 0,82 | 10.0 [4.3-15.0] | 10.0 [5.0-12.5] | 0,95 |
| IT TILs (mean) |  | 10.0 [5.0-12.5] | 5.0 [3.0-5.0] | 0,02 | 5.0 [1.8-6.3] | 5.0 [3.0-10.0] | 0,54 | 5.0 [5.0-5.0] | 5.0 [2.0-5.0] | 0,68 |

Missing data: Menopausal status, n=1; BMI (continuous), n=1; BMI class, n=1; Family history, n=3; Grade, n=5; Mitotic index, n=77; Pre-NAC str TILs, n=75; Pre-NAC IT TILs, n=75; NAC regimen, n=1; pCR status, n=1; Post-NAC Nodal involvement, n=1; Post-NAC str TILs, n=75; Post-NAC IT TILs, n=147.
NAC=neoadjuvant chemotherapy; BMI=body mass index; NST= no special type; TNBC= triple negative breast cancer; str TILs= stromal tumor-infiltrating lymphocytes; IT TILs= intratumoral-infiltrating lymphocytes; AC=anthracyclines; pCR=Pathologic complete response.
The "n" denotes the number of patients. In case of categorical variables, percentages are expressed between brackets. In case of continuous variables, mean value is reported. In case of nonnormal continuous variables, median value is reported, with interquartile range between brackets.

**Supplementary Table S3.** Association of BRCA status with pCR after univariate and multivariate analysis in the whole population

|  |  |  |  |  | Univariate |  |  |  | Multivariate |  |  |
| --- | --- | --- | --- | --- | --- | --- | --- | --- | --- | --- | --- |
| Variable | Class | Nb total | Nb in model | Events | HR | CI | RCH | p | HR | CI | p |
| Pre-NAC parameters |  |  |  |  |  |  |  |  |  |  |  |
| Age (years) |  |  |  |  |  |  |  |  |  |  |  |
| Menopausal status | pre | 227 | 227 | 70 | 1 |  | 30.8 % |  |  |  |  |
|  | post | 38 | 38 | 14 | 1.31 | [0.63 - 2.65] | 36.8 % | 0,46 |  |  |  |
| BMI class | ≤19 | 23 | 23 | 8 | 1 |  | 34.8 % |  |  |  |  |
|  | 19-25 | 177 | 177 | 56 | 0.87 | [0.36 - 2.27] | 31.6 % | 0,76 |  |  |  |
|  | 25-30 | 47 | 47 | 17 | 1.06 | [0.38 - 3.11] | 36.2 % | 0,90 |  |  |  |
|  | >30 | 18 | 18 | 3 | 0.37 | [0.07 - 1.58] | 16.7 % | 0,20 |  |  |  |
| BRCA status | BRCA mutation | 46 | 46 | 21 | 1 |  | 45.7 % |  |  |  |  |
|  | BRCA wild-type | 220 | 220 | 63 | 0.48 | [0.25 - 0.92] | 28.6 % | **0,03** |  |  |  |
| Tumor size | T1 | 27 | 27 | 15 | 1 |  | 55.6 % |  |  |  |  |
|  | T2 | 181 | 181 | 57 | 0.37 | [0.16 - 0.83] | 31.5 % | **0,02** | 0.51 | [0.17 - 1.49] | 0.22 |
|  | T3 | 58 | 58 | 12 | 0.21 | [0.08 - 0.55] | 20.7 % | **<0.01** | 0.26 | [0.07 - 0.89] | **0.03** |
| Clinical nodal status | N0 | 110 | 110 | 33 | 1 |  | 30% |  |  |  |  |
|  | N1 | 145 | 145 | 48 | 1.15 | [0.68 - 1.98] | 33.1 % | 0,61 |  |  |  |
|  | N2 | 8 | 8 | 2 | 0.78 | [0.11 - 3.58] | 25% | 0,77 |  |  |  |
|  | N3 | 3 | 3 | 1 | 1.17 | [0.05 - 12.59] | 33.3 % | 0,91 |  |  |  |
| Grade | I | 7 | 7 | 1 | 1 |  | 14.3 % |  |  |  |  |
|  | II | 78 | 78 | 17 | 1.67 | [0.26 - 32.72] | 21.8 % | 0,65 |  |  |  |
|  | III | 176 | 176 | 66 | 3.6 | [0.6 - 68.78] | 37.5 % | 0.24 |  |  |  |
| Mitotic index | ≤22 | 101 | 101 | 29 | 1 |  | 28.7 % |  |  |  |  |
|  | >22 | 88 | 88 | 34 | 1.56 | [0.85 - 2.89] | 38.6 % | 0.15 | 0.89 | [0.42 - 1.83] | 0.74 |
| BC subtype | luminal | 89 | 89 | 9 | 1 |  | 10.1 % |  |  |  |  |
|  | TNBC | 110 | 110 | 49 | 7.14 | [3.39 - 16.57] | 44.5 % | **<0.01** | 8.6 | [2.76 - 33.41] | **<0.01** |
|  | HER2 | 67 | 67 | 26 | 5.64 | [2.5 - 13.78] | 38.8 % | **<0.01** | 6.31 | [1.99 - 24.48] | **<0.01** |
| str TILs (%) |  |  |  | 84 | 1.03 | [1.02 - 1.05] |  | **<0.01** | 1.01 | [0.99 - 1.04] | 0.33 |
| IT TILs (%) |  |  |  | 84 | 1.04 | [1.02 - 1.07] |  | **<0.01** | 1.03 | [0.98 - 1.07] | 0.24 |
| NAC regimen | AC | 29 | 29 | 7 | 1 |  | 24.1 % |  |  |  |  |
|  | AC-Taxanes | 221 | 221 | 72 | 1.52 | [0.65 - 3.99] | 32.6 % | 0,36 |  |  |  |
|  | Taxanes | 7 | 7 | 3 | 2.36 | [0.39 - 13.48] | 42.9 % | 0,33 |  |  |  |
|  | Others | 9 | 9 | 2 | 0.9 | [0.12 - 4.86] | 22.2 % | 0,91 |  |  |  |

Abbreviation, BMI=body mass index; ER=oestrogen receptor; PR=progesterone receptor; TNBC= triple negative breast cancer; str TILs= stromal tumor-infiltrating lymphocytes ; IT TILs= intratumoral-infiltrating lymphocytes ; NAC=neoadjuvant chemotherapy ; AC=anthracyclines ; pCR=Pathologic complete response.

## Supplementary Figures

**Supplementary Figure S 1.**
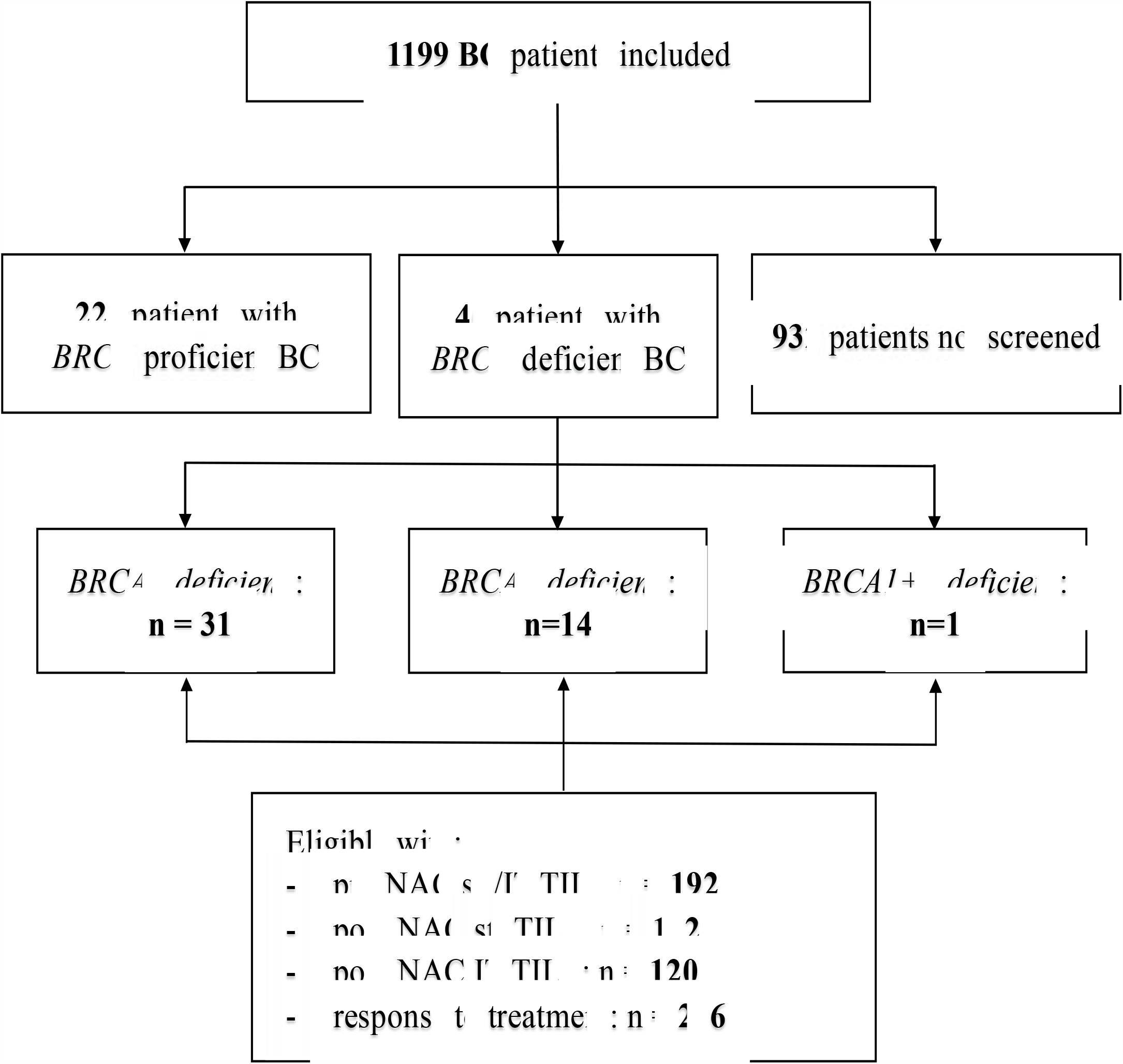
Study flow diagram of included patients and tumors samples available

**Supplementary Figure S 2.**
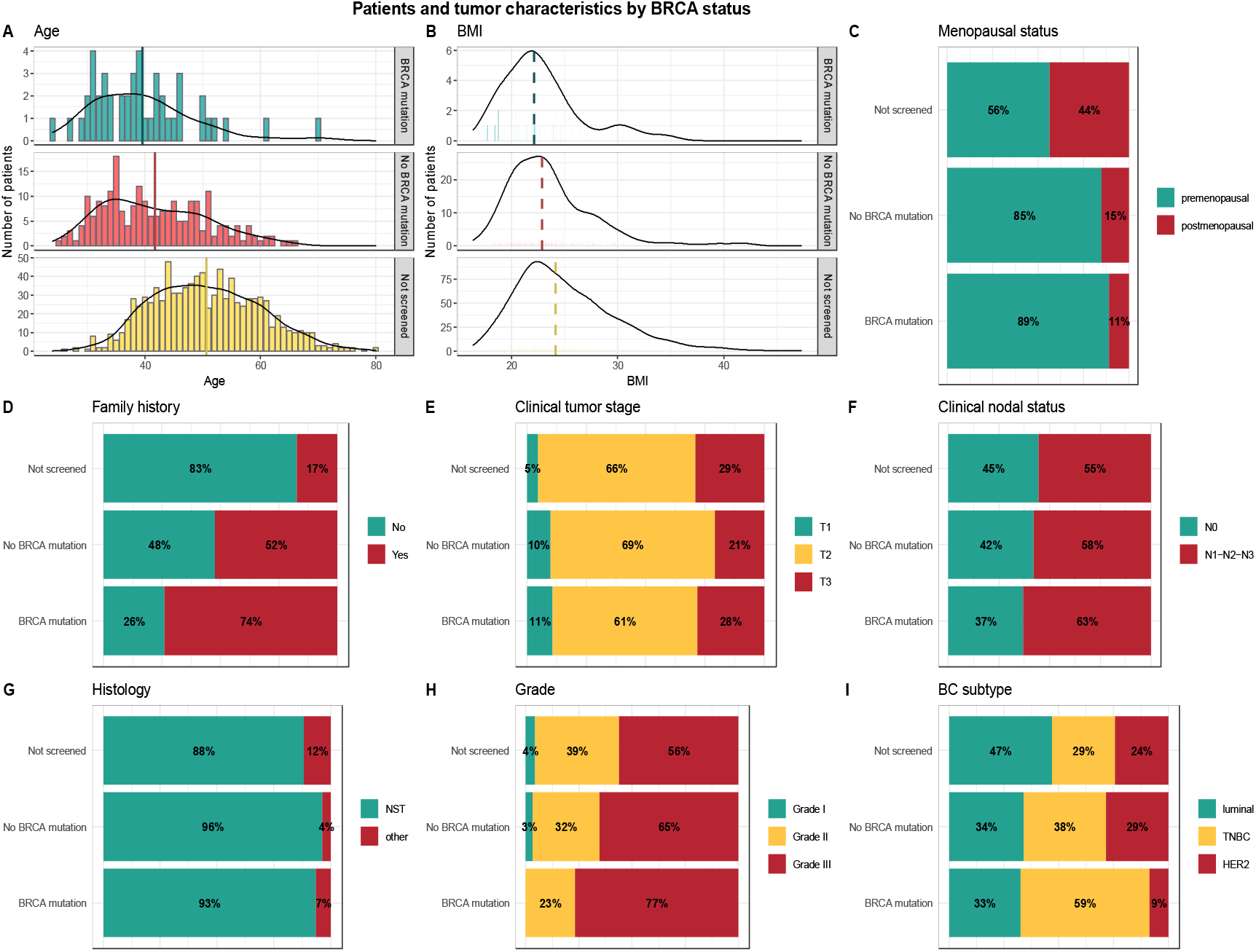
Patients’ and tumors ‘characteristics by BRCA status. (All(n=1199), BRCA mutation (n=36), BRCA wild-type(n=156), not screened (n=1007). A, Age (kernel density plot). B, BMI (kernel density plot). C, Menopausal status (barplot). D, Family history (barplot). E, Clinical tumor stage (barplot). F, Clinical nodal status (barplot). G, Histology (barplot). H, Grade (barplot). I, BC subtype (barplot).

**Supplementary Figure S 3.**
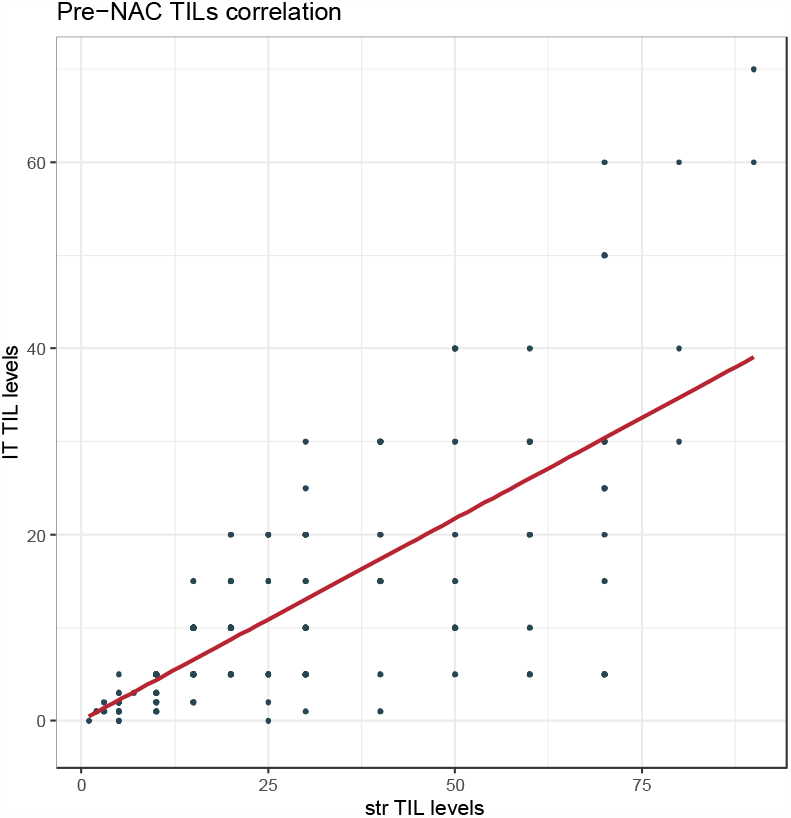
Variation of pre-NAC str TIL levels according to the pre-NAC IT TIL levels (scatterplot) (str TILs (n=192), IT TILs (n=192)).

**Supplementary Figure S 4.**
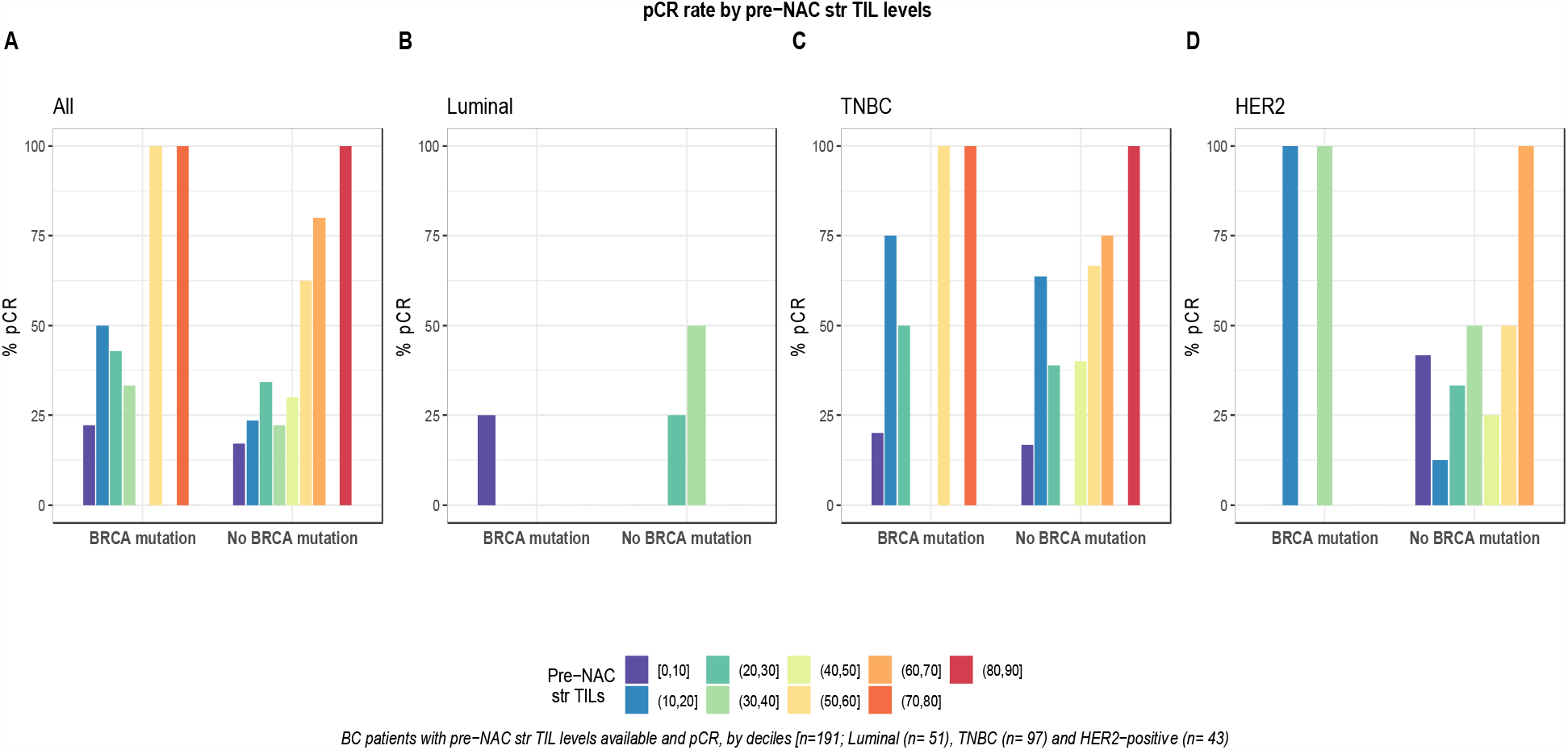
pCR rate by pre-NAC str TIL levels by BRCA status (TILs were binned by increments of 10%). A, whole population (n=191, BRCA mutation (n=36), BRCA wild-type(n=155)). B, luminal tumors (n=51, BRCA mutation(n=8), BRCA wild-type(n=43)). C, TNBC (n=97), BRCA mutation(n=24), BRCA wild-type(n=73)). D, HER2-positive BC (n=43, BRCA mutation(n=4), BRCA wild-type(n=39)).

**Supplementary Figure S 5.**
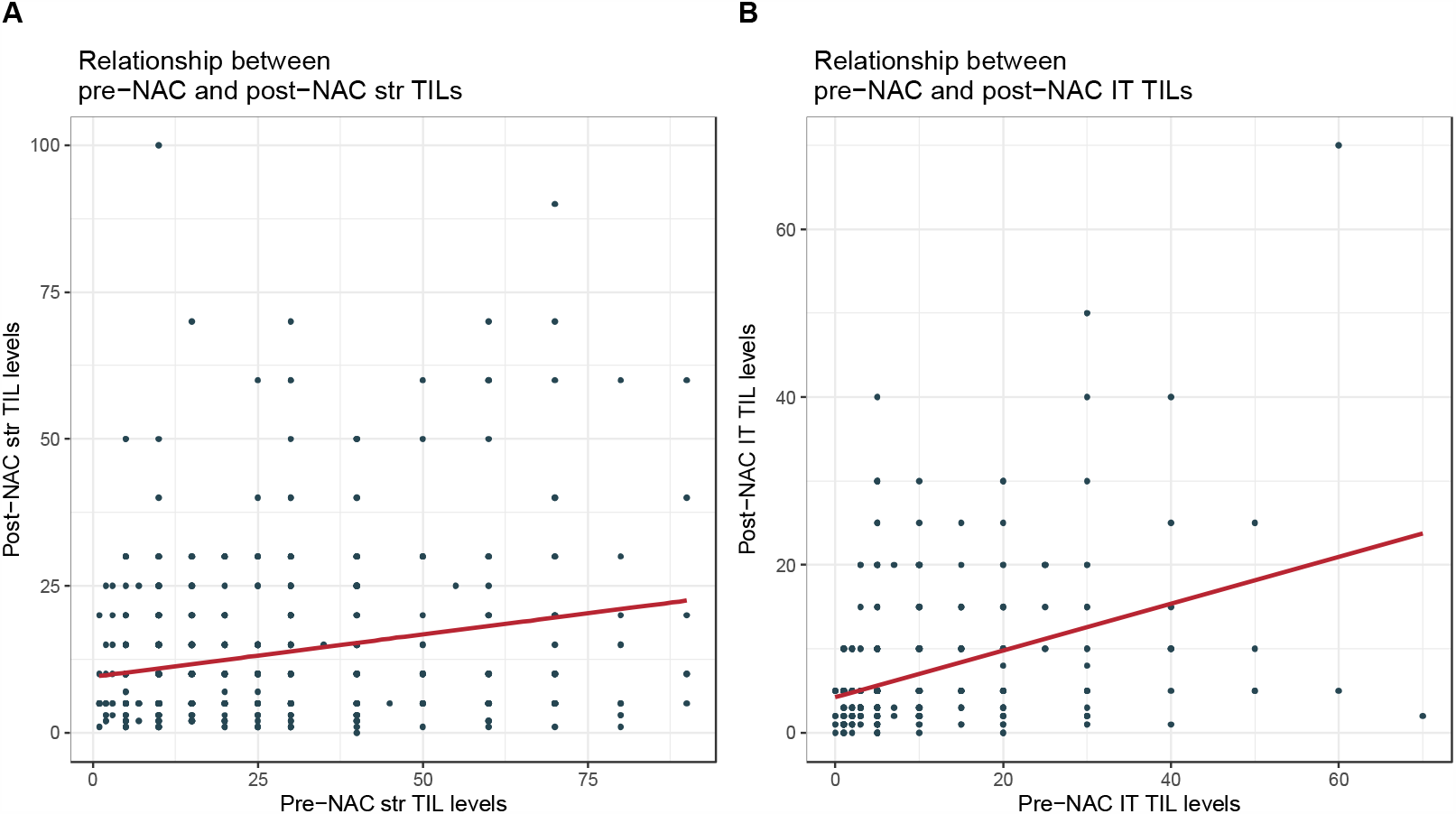
TILs correlation between pre and post-NAC. A, Variation of post-NAC str TIL levels according to the pre-NAC str TIL levels (scatterplot) (pre-NAC str TILs (n=192), post-NAC str TILs (n=192)). B, Variation of post-NAC IT TIL levels according to the pre-NAC IT TIL levels (scatterplot) (pre-NAC IT TILs (n=192), post-NAC IT TILs (n=120)).

**Supplementary Figure S 6.**
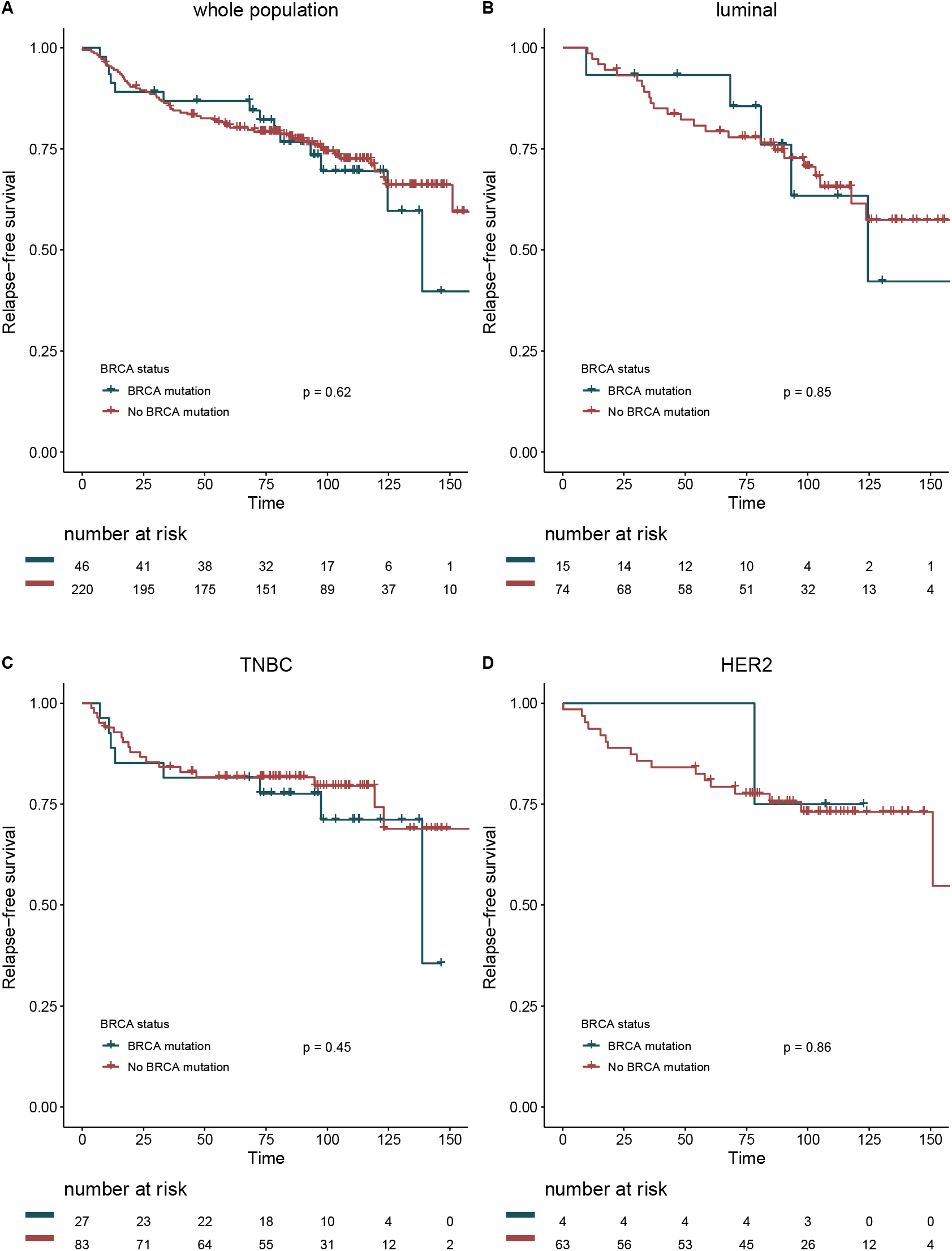
Relapse free survival curves according BRCA status. A, whole population (n=267, BRCA mutation (n=46), BRCA wild-type(n=220)). B, luminal tumors (n=89, BRCA mutation(n=15), BRCA wild-type(n=74)). C, TNBC (n=110, BRCA mutation(n=27), BRCA wild-type(n=83)). D, HER2-positive BC (n=67), BRCA mutation(n=4), BRCA wild-type(n=63)).

**Supplementary Figure S 7.**
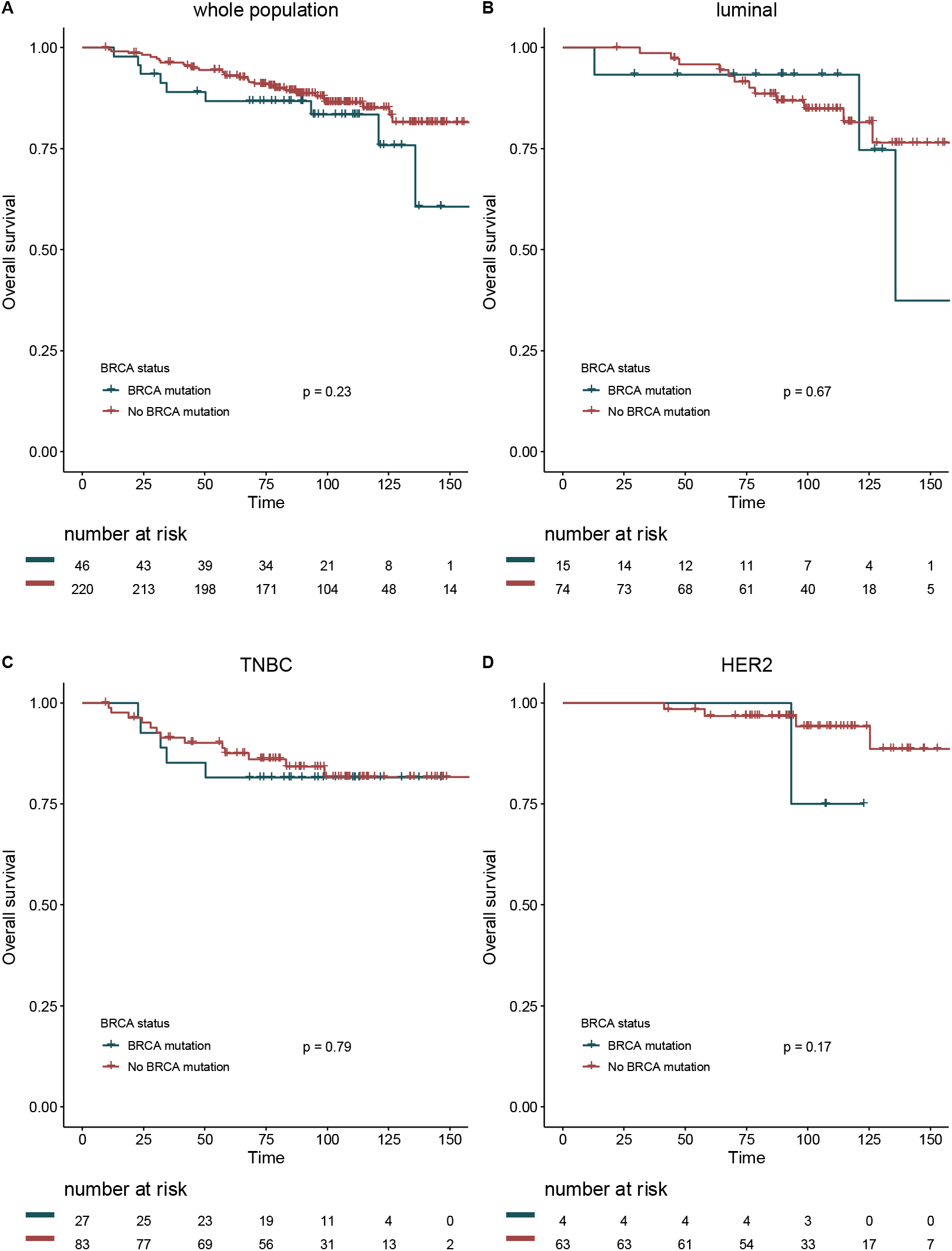
Overall survival curves according BRCA status. A, whole population (n=267, BRCA mutation (n=46), BRCA wild-type(n=220)). B, luminal tumors (n=89, BRCA mutation(n=15), BRCA wild-type(n=74)). C, TNBC (n=110, BRCA mutation(n=27), BRCA wild-type(n=83)). D, HER2-positive BC (n=67), BRCA mutation(n=4), BRCA wild-type(n=63)).

## Notes

**Financial Support:** B.G.R. was supported by Alfonso Martin Escudero Foundation research grant.

### Competing Interest Statement

The authors have declared no competing interest.

### Funding Statement

Beatriz Grandal was supported by Alfonso Martin Escudero Foundation research grant.

### Author Declarations

Approved by the Breast Cancer Study Group of Institute Curie, the study was conducted according to institutional and ethical rules concerning research on tissue specimens and patients. Informed consent from patients was not required.

